# State-level structural racism and incident coronary heart disease

**DOI:** 10.1101/2024.11.01.24316616

**Authors:** Monika M. Safford, Tyson Brown, Joanna Bryan, Todd M. Brown, Laura Pinheiro

**Affiliations:** Division of General Internal Medicine, Weill Medical College of Cornell University, New York, NY; Departments of Sociology and Medicine, Duke University, Durham, NC; Division of Cardiovascular Disease, University of Alabama at Birmingham Heersink School of Medicine, Birmingham, AL

**Keywords:** structural racism, incidence coronary heart disease, racial disparities, cohort studies

## Abstract

**Introduction:** Black Americans have greater coronary heart disease (CHD) burden than White Americans, disparities that are largely socially determined. Discriminatory societal practices that systematically disadvantage Black Americans are forms of structural racism but few studies have examined structural racism and incident CHD. We sought to determine associations between three validated measures of structural racism and incident CHD, hypothesizing that greater state-level structural racism is associated with incident CHD for Black but not White individuals.

**Methods:** We used data from the national REasons for Geographic And Racial Differences in Stroke (REGARDS) cohort, which enrolled 30,239 Black and White community dwelling adults between 2003-7 who were contacted every 6 months with retrieval of medical records and expert adjudication of myocardial infarction and cause of death. Incident CHD was defined as myocardial infarction or death due to CHD. Structural racism variables included Black:White % living below the Federal poverty line, Black:White % uninsured, and the Dissimilarity Index (DI), a measure of residential racial segregation. Structural racism variables were dichotomized at the median. Separate race-stratified Cox proportional hazards models examined associations between each measure of structural racism and incident CHD.

**Results:** The 24,099 participants free of CHD at baseline included 10,286 Black and 13,813 White participants. Mean age at baseline was 64 years, 58% were women, and 47% had annual household income <$35,000. Greater structural racism was significantly associated with incident CHD for Black but not White participants. For high Black:White poverty, Black HR=1.17 (95% CI 1.01-1.35), White HR=0.93 (0.83-1.06); for high Black:White uninsurance, Black=HR 1.34 (1.06-1.70), White HR=1.20 (0.98-1.47); for high DI, Black HR=1.17 (1.01-1.35), White HR=0.99 (0.88-1.12). Findings suggest that structural racism variables indirectly influence CHD via individual-level income and education. Results were similar for men and women and for older and younger individuals. Significant associations were observed for fatal but not nonfatal CHD events.

**Conclusions:** Structural racism was associated with higher incidence of CHD for Black but not White individuals. If these associations are causal, changing state level laws to combat poverty in Black communities, expand Medicaid, and reduce segregation could potentially lessen Black:White disparities in CHD.

## Introduction

Coronary heart disease (CHD) inequities along racial lines in the US are long standing and pervasive.^1^ A large body of work has described the persistence of CHD racial inequities, but few studies have examined potential upstream root causes that could be modified by policy change at the state level. A small but growing body of evidence supports the notion that the way we have chosen to structure our societal institutions may have a large influence on racial health inequities.^2–4^ For example, discriminatory practices undergirding systematic disadvantages for Black people vis-à-vis their White counterparts across societal domains—e.g., economy, health care, and housing^5,6,7^— has resulted in Black people experiencing higher rates of poverty and uninsurance as well as residential segregation; these have been shown to be powerful risk factors for poor health outcomes, including CHD.^8,9,10^ In the present era, low income is also associated with lack of health insurance, since many people with low income are restricted to jobs that do not offer employer-based health insurance. However, the upstream societal structures that have restricted economic opportunity for Black individuals and lead to high poverty have rarely been studied in relation to incident CHD. Investigating such associations is useful for informing efficacious policy efforts aimed at achieving more equitable health outcomes.

We sought to fill this knowledge gap by examining area level variables that reflect structural racism and test their association with incident fatal and nonfatal CHD. Since many policies that influence residential segregation and restrict access to economic opportunity and healthcare are set at the state level, we operationalized structural variables at the state level rather than at the census tract.^11–13^ We hypothesized that Black REGARDS participants residing in a State with above median: 1) Black-to-White poverty ratio; 2) Black-to-White ratio of uninsured individuals, and 3) residential racial segregation as reflected in the Dissimilarity Index (DI); would have higher rates of incident CHD, but White REGARDS participants residing in the same State would not. By investigating the relationship between structural racism and risk of incident CHD in the REGARDS cohort, we aimed to shed light on the role of structural racism in risk for incident CHD for Black and White individuals in the US.

## Methods

### Dataset

The REasons for Geographic And Racial Differences in Stroke (REGARDS) study (IRB: 1603017100) was originally designed to identify and understand reasons for regional and racial differences in stroke mortality; CHD outcomes are being investigated through an ancillary study (R01HL80477 and R01HL165452). From 2003-2007, community-dwelling self-identified Black and White adults aged 45 years or older living in the 48 contiguous US states were consented and enrolled in accordance with institutional guidelines. The sampling strategy aimed to balance race and sex, and purposely oversampled the regions known as the Stroke Buckle (coastal North and South Carolina and Georgia) and the Stroke Belt (the remainder of North and South Carolina and Georgia, and Alabama, Mississippi, Louisiana, Arkansas, and Tennessee), which have higher stroke mortality than the rest of the country. Participants were recruited using commercially available lists and underwent 45-minute computer assisted telephone interviews (CATI) about medical history, health behaviors, and risk factors.

Interviews were followed by in-home visits at which physiologic parameters (height, weight, blood pressure) and electrocardiography were assessed. Blood and urine samples were collected and shipped to the study’s central laboratory at the University of Vermont. An inventory of medications taken in the two weeks prior to the study visit was obtained through pill bottle review, and a food frequency questionnaire was left for self-administration. Participants were contacted every 6 months by telephone to detect potential study endpoints.

### Study sample and ethical standards

We included participants who were free of CHD at baseline, defined as self-reported history of myocardial infarction (MI), coronary angioplasty or stenting, coronary artery bypass graft; or old MI on the baseline ECG. The institutional review boards of the participating institutions approved the study, and all participants provided written informed consent.

### Primary outcome

This study’s primary outcome was expert adjudicated incident CHD, defined as acute MI or fatal CHD. Hospitalizations for potential study outcomes detected during follow-up triggered medical record retrieval and adjudication by clinician experts using national guidelines.^14,15^ Acute MI was classified based on clinical presentation, a rise and/or fall in biomarkers (troponin), and electrocardiographic findings.^16^ Fatal CHD was defined as sudden death, death within 28 days of an adjudicated definite or probable MI, or death in a participant with known CHD and no other non-coronary cause of death. The main underlying cause of death was determined by two trained adjudicators who examining all available information including interviews with next of kin, death certificates, autopsy reports, medical history, and the National Death Index.

Disagreements between the two reviewers were resolved by committee. Events through December 31, 2020 were available.

### Key explanatory variables

We considered three validated measures of structural racism at the state level. These included Black:White % living below the Federal poverty line in the state,^13^ Black:White % uninsured in the state,^12^ and the Dissimilarity Index (DI), which reflects the proportion of Black individuals who would have to move to achieve a similar demographic composition as a state’s proportion of Black residents in all areas in that state.^11^ The DI ranges from 0 to 1 with higher scores indicating that a greater proportion of Black residents would need to move, or a higher degree of residential segregation. Poverty and insurance were derived from the 2010 US census. The DI was constructed from 2010 data obtained from the National Strategic Planning and Analysis Residential Segregation Research Center. Structural racism variables were dichotomized at the median.^12^

### Covariates

We controlled for three variables collected at the baseline REGARDS survey: region (stroke belt, buckle, non-belt/buckle), gender, and age. At the state level, we considered the percent of Black individuals residing in the state and the overall rates of state level poverty and uninsurance. To better understand potential pathways through which structural racism influences risk of CHD, we also used a nested model strategy that adds groups of variables representing individual social position (income and education), cardiovascular disease (CVD) risk factors (hypertension, diabetes, c-reactive protein, albumin-to-creatinine ratio, hyperlipidemia), psychosocial variables (depressive symptoms, loneliness, perceived stress), and health behaviors (cigarette smoking, alcohol consumption).

### Statistics

We first calculated descriptive statistics for study participants by race (Black vs. White). Chi square tests were used to determine statistically significant differences for categorical covariates and ANOVA or Kruskal Wallis tests were applied for continuous covariates. We then examined the distribution of each measure of structural racism to identify the median value in our sample, which was used to dichotomize each measure. That is, for each measure, participants were classified as either living in a state at or above or below the median for the variable in question.

To examine race-specific relationships between structural racism measures and CHD risk, all models were stratified by race. Cox proportional hazard models with robust standard errors were used to estimate the association between each structural racism measure and incident CHD, separately for Black and White participants. We calculated minimally adjusted hazard ratios (HR) and 95% confidence intervals (95% CI) controlling for age, gender, and region. We then included the % of Blacks within state to account for the racial composition, and the overall rates of poverty and uninsurance.

Next, socioeconomic status, CHD risk factors, psychosocial factors, and health behaviors were added individually and then removed from the model, observing changes in magnitude and directionality and statistical significance in the HR for the structural racism variable. Finally, we estimated fully adjusted HR controlling for all covariates in the model simultaneously. For all estimates, 95% CI were calculated and p-values < 0.05 were considered statistically significant.

To understand differences in the risk potentially incurred for fatal vs. nonfatal incident CHD events, we repeated the above analyses modeling each CHD outcome separately. Because Black women are uniquely burdened by health inequities and poverty, we examined interactions between each structural factor and sex within each race stratum. Our own prior work in REGARDS as well as the work of others suggest that disparities are worst at younger ages; therefore, we also examined interactions between the structural variables and age using a Wald test of interaction terms for each structural racism measure with gender (male vs. female) and age (<65 years vs. 65+ years). P-values below 0.10 were considered indicative of potential effect modification.

Bias due to missing information of covariates was mitigated using multiple imputation with chained equations. Results were summarized using Rubin’s rules across 20 imputations.

All analyses were conducted in SAS, version 9.4 (SAS Institute) and Stata, version 17 (StataCorp LLC). Dr. Monika M Safford had full access to all the data in the study and takes responsibility for its integrity and the data analysis.

## Results

### Study participants

Our analytic cohort included 24,099 participants free of CHD at baseline. Of these, 10,286 (42.7%) self-identified as Black and 13,813 (57.3%) self-identified as White. The mean age at baseline was 64 (SD 9.3 years, 58.5% were women, and 47% had annual household income <$35,000 (Table 1).

**Figure 1.**
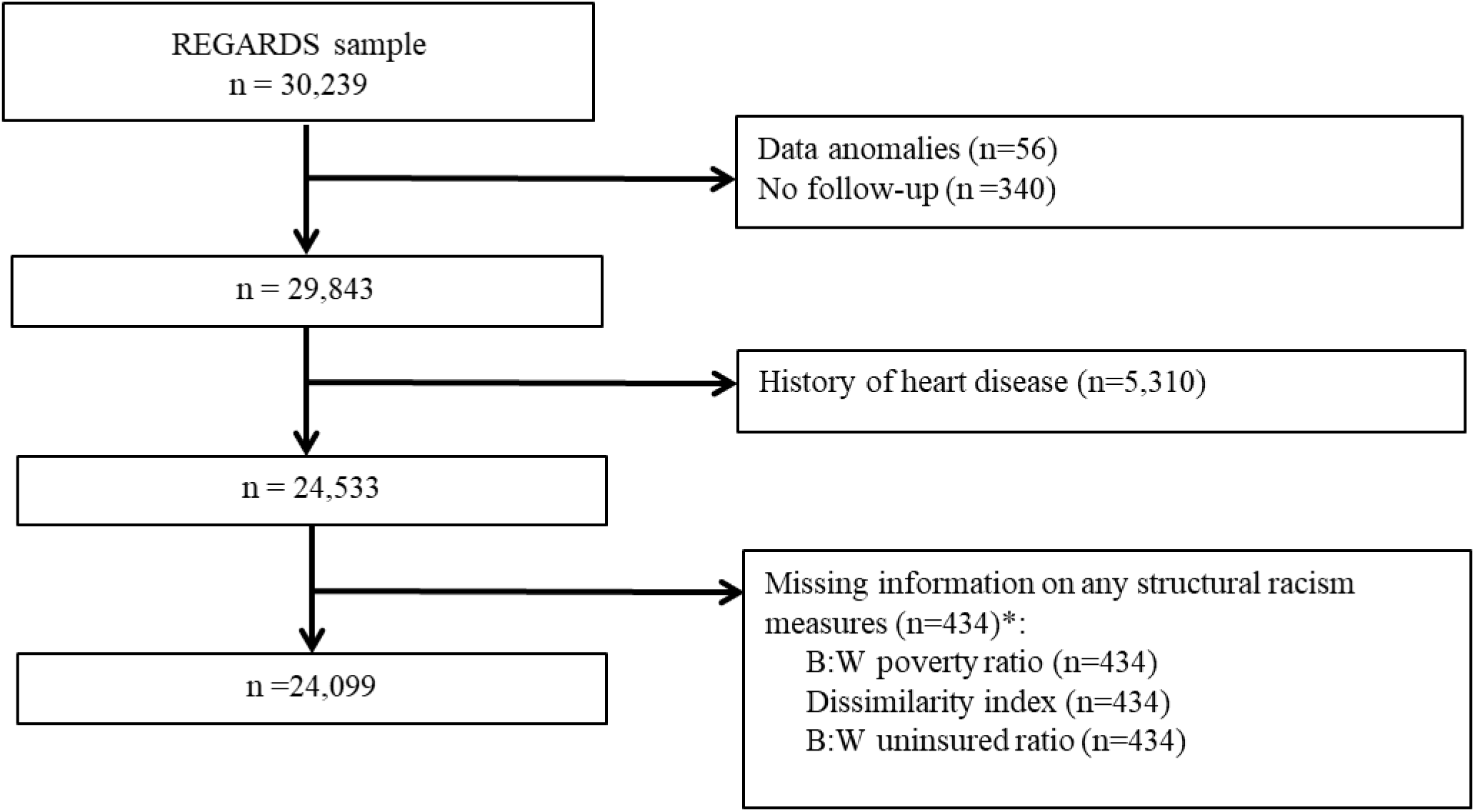
Exclusion cascade for the study. Abbreviations: REGARDS- Reasons for geographic and racial differences in strokes *Participants living in the District of Columbia and the following states were excluded from the analysis because administrative data on Black people was not available due to their very small population size: Idaho, Maine, Montana, New Hampshire, New Mexico, North Dakota, Oregon, South Dakota, Utah, Vermont, Wyoming

**Table 1.** Characteristics of participants without history of heart disease at baseline, overall and by race.

| Factor | Total | Black | White | P-value |
| --- | --- | --- | --- | --- |
| N | 24099 | 10286 | 13813 |  |
| **Demographics** |  |  |  |  |
| Age, mean (SD) | 64.1 (9.33) | 63.6 (9.27) | 64.5 (9.36) | <0.001 |
| Women, n (%) | 14093 (58.5%) | 6559 (63.8%) | 7534 (54.5%) | <0.001 |
| Region, n (%) |  |  |  | <0.001 |
| Non-belt | 10468 (43.4%) | 3515 (34.2%) | 4992 (36.1%) |  |
| Belt | 8507 (35.3%) | 5370 (59.7%) | 4511 (37.3%) |  |
| Buckle | 5124 (21.3%) | 1883 (18.3%) | 3241 (23.5%) |  |
| **Social position** |  |  |  |  |
| Income < \$35,000, n (%) | 9881 (46.8%) | 4888 (47.5%) | 5580 (40.4%) | <0.001 |
| Low educational attainment, n (%) | 2826 (11.7%) | 1945 (18.9%) | 881 (6.4%) | <0.001 |
| **CVD risk factors** |  |  |  |  |
| Hypertension, n (%) | 13590 (56.5%) | 7115 (69.3%) | 6475 (47.0%) | <0.001 |
| Diabetes, n (%) | 4569 (19.7%) | 2832 (28.8%) | 1737 (13.0%) | <0.001 |
| C-Reactive protein (mg/L), median (IQR) | 2.20 (0.95, 4.98) | 2.85 (1.17, 6.39) | 1.84 (0.83, 4.14) | <0.001 |
| Albumin to creatinine ratio >30 mg/dL, n (%) | 3098 (13.5%) | 1722 (17.7%) | 1376 (10.4%) | <0.001 |
| Hyperlipidemia, n (%) | 12866 (55.6%) | 5141 (52.6%) | 7725 (57.8%) | <0.001 |
| PCS-12, median (IQR) | 50.6 (41.6, 55.2) | 49.2 (39.2, 54.0) | 51.7 (43.3, 55.5) | <0.001 |
| **Psychosocial factors** |  |  |  |  |
| Perceived Stress Scale, median (IQR) | 3.0 (0.00, 5.00) | 3.0 (1.00, 6.00) | 2.0 (0.00, 5.00) | <0.001 |
| Depressive symptoms, n (%) | 2517 (10.5%) | 1347 (13.2%) | 1170 (8.5%) | <0.001 |
| Social isolation (0-1 friends/family seen during past month), n (%) | 2931 (12.4%) | 1420 (14.1%) | 1511 (11.1%) | <0.001 |
| Social isolation (someone to provide care when ill), n (%) | 3162 (14.1%) | 1361 (14.3%) | 1801 (13.9%) | 0.34 |
| MCS-12, median (IQR) | 56.7 (51.7, 59.3) | 56.4 (50.3, 59.4) | 56.8 (52.7, 59.2) | <0.001 |
| Feeling lonely ≥1 day per week, n (%) | 4833 (20.4%) | 2565 (25.4%) | 2268 (16.7%) | <0.001 |
| **Health behaviors** |  |  |  |  |
| Current cigarette smoking, n (%) | 3418 (14.2%) | 1731 (16.9%) | 1687 (12.3%) | <0.001 |
| Exercising enough to work up a sweat at least once/week, n (%) | 15764 (66.4%) | 6431 (63.4%) | 9333 (68.6%) | <0.001 |
| Alcohol use, n (%) |  |  |  | <0.001 |
| None | 14752 (62.4%) | 7239 (72.2%) | 7513 (55.2%) |  |
| Moderate | 7882 (33.4%) | 2544 (25.4%) | 5338 (39.2%) |  |
| Heavy | 1000 (4.2%) | 245 (2.4%) | 755 (5.5%) |  |

Abbreviations: CVD = cardiovascular disease, MCS = mental component summary score of the Short Form 12-item questionnaire, PCS =
physical component summary score of the Short Form 12-item questionnaire, SD = standard deviation.

### Incident CHD

Over a median follow-up of 13.0 years (IQR 7.0-13.0), we observed 1,926 incident CHD events (1,247 nonfatal and 679 fatal), 800 (7.8%) among Black participants and 1,126 (8.1%) among White participants (p = 0.29).

### Structural racism and incident CHD

Greater structural racism at the state level was statistically significantly associated with incident CHD for Black but not White participants (Table 2). These patterns were observed for all three measures of structural racism.

- *Poverty:* For high Black:White poverty, adjusting for age, sex, and region, the HR was 1.19 (95% CI 1.03-1.37) for Black participants and the HR was 0.90 (95% CI 0.80-1.01) for White participants. After adjusting for the percent of Black residents in the state and the overall rate of individuals living below the federal poverty line in the state, the HR for Black participants attenuated slightly (Black HR 1.17 [95% CI 1.01-1.35], White HR 0.93 [95% CI 0.83-1.06]).
- *Lack of health insurance:* For high Black:White uninsurance, adjusting for age, sex, and region, the HR was 1.18 (95% CI 1.02-1.36) for Black participants and the HR was 1.01 (95% CI 0.89-1.14) for White participants. After adjusting for the percent of Black residents in the state and the overall uninsurance rate of individuals in the state, the HR increased to 1.34 (95% CI 1.06-1.70) for Black participants and the HR was 1.20 (95% CI 0.98-1.47) for White participants.
- *Dissimilarity Index:* For high DI, adjusting for age, sex, and region, the HR was 1.16 (95% CI 1.01-1.34) for Black participants and the HR was 0.99 (95% CI 0.88-1.12) for White participants. After adjusting for the percent of Black residents in the state, the HR was 1.17 (95% CI 1.01-1.35) for Black participants and the HR remained at 0.99 (0.88-1.12) for White participants.

**Table 2.** Hazard ratios for the association between structural racism measures and incident CHD.

| Structural Racism Measure | Model 1 |  | Model 2 |  | Model 3 |  | Model 4 |  | Model 5 |  | Model 6 |  |
| --- | --- | --- | --- | --- | --- | --- | --- | --- | --- | --- | --- | --- |
|  | HR (95% CI) | p | HR (95% CI) | p | HR (95% CI) | p | HR (95% CI) | p | HR (95% CI) | p | HR (95% CI) | p |
| **Black participants** |  |  |  |  |  |  |  |  |  |  |  |  |
| High B:W % poverty | 1.17<br>(1.01,1.35) | 0.04 | 1.11<br>(0.96,1.28) | 0.18 | 1.13<br>(0.98,1.31) | 0.10 | 1.15<br>(0.99,1.33) | 0.06 | 1.15<br>(0.99,1.33) | 0.06 | 1.10<br>(0.95,1.28) | 0.20 |
| High B:W uninsured rate | 1.34<br>(1.06,1.70) | 0.02 | 1.26<br>(0.99,1.60) | 0.06 | 1.28<br>(1.01,1.63) | 0.04 | 1.33<br>(1.05,1.68) | 0.02 | 1.31<br>(1.04,1.66) | 0.02 | 1.23<br>(0.97,1.56) | 0.09 |
| High dissimilarity index | 1.17<br>(1.01,1.35) | 0.03 | 1.13<br>(0.97,1.30) | 0.11 | 1.15<br>(0.99,1.33) | 0.06 | 1.16<br>(1.01,1.35) | 0.04 | 1.15<br>(1.00,1.34) | 0.05 | 1.12<br>(0.96,1.29) | 0.14 |
| **White participants** |  |  |  |  |  |  |  |  |  |  |  |  |
| High B:W % poverty | 0.93<br>(0.83,1.06) | 0.28 | 0.92<br>(0.82,1.04) | 0.21 | 0.91<br>(0.80,1.03) | 0.13 | 0.93<br>(0.82,1.06) | 0.27 | 0.92<br>(0.81,1.04) | 0.17 | 0.89<br>(0.79,1.01) | 0.07 |
| High B:W uninsured rate | 1.20<br>(0.98,1.47) | 0.07 | 1.19<br>(0.97,1.45) | 0.09 | 1.17<br>(0.96,1.43) | 0.12 | 1.20<br>(0.98,1.47) | 0.07 | 1.17<br>(0.95,1.43) | 0.13 | 1.16<br>(0.95,1.42) | 0.16 |
| High dissimilarity index | 0.99<br>(0.88,1.12) | 0.90 | 0.98<br>(0.87,1.10) | 0.72 | 0.97<br>(0.86,1.10) | 0.65 | 0.99<br>(0.88,1.12) | 0.89 | 0.97<br>(0.86,1.09) | 0.59 | 0.95<br>(0.85,1.08) | 0.44 |

Model 1 adjusts for age, gender, region, % Black residents in the state, and the overall rate of the variable for all residents of the state for poverty and uninsurance
Model 2 adjusts for model 1 covariates plus individual income and education
Model 3 adjusts for model 1 covariates plus CVD risk factors (hypertension, diabetes, log transformed C reactive protein, albumin-to-creatinine ratio > 30, hyperlipidemia)
Model 4 adjusts for model 1 covariates plus psychosocial risk factors (depression, feeling lonely, perceived stress)
Model 5 adjusts for model 1 covariates plus behavioral risk factors (smoking and alcohol)
Model 6 adjust for all variables
For each structural racism variable, participants were classified as either living in a state at or above vs. below median values for the variable in the overall REGARDS sample.

Adding covariates to the adjusted models described above revealed that the strongest potential mediators of the relationships described above were individual annual household income and highest level of education (Table 2). Among Black participants, adding income and education to the models adjusting for age, gender, region, % Black residents, and the overall rate of the variable being studied slightly attenuated the HRs for Black:White poverty rates and segregation and they became nonsignificant, with remaining borderline significance for Black:White uninsurance rates. No substantive changes occurred for White participants. In contrast, adding CVD risk factors, psychosocial risk factors, or behavioral risk factors to the base models attenuated the HR less than the addition of individual income and education, and the elevated risk remained statistically significant for Black participants with little change for White participants. Among Black participants, the HR attenuated with all covariates added to the models and became nonsignificant for all three structural racism variables.

Findings were similar for men and women and for older and younger individuals, i.e., none of the tests for interaction had p <0.10.

The analyses examining fatal and nonfatal CHD outcomes separately showed that the associations between the 3 structural racism factors and incident CHD events were present for fatal but not nonfatal CHD events (supplemental Tables 1 and 2). For fatal CHD, among Black participants, the HR for B:W % living below the federal poverty line was 1.30 (95% CI 1.03-1.63); the HR for B:W uninsurance was 1.77 (95% CI 1.23- 2.55); and the HR for the DI was 1.32 (95% CI 1.04-1.66). The largest contribution to these observed risks was individual income and education. Notably, the HR for uninsurance remained significant even after entering all explanatory variables into the model for incident fatal CHD. There were no significant associations for White participants except for uninsurance for incident fatal CHD; the HR was 1.42 (95% CI 1.01-2.00) for the base model and this attenuated to 1.37 (95% CI 0.98-1.93) with full adjustment.

## Discussion

In a national, longitudinal cohort of more than 24,000 Americans, three state-level structural racism domains were significantly associated with higher incidence of CHD over 13 years for Black but not White individuals. This risk was demonstrable for incident fatal CHD but not incident nonfatal CHD. Our findings are consistent with the notion that structural factors in our society are root causes of racial inequities that could be intervened upon to achieve equity in CHD outcomes.

Our study answers recent calls by several leaders in the cardiovascular research community to address structural racism directly. Churchwell, et al, recommended that research on the health effects of structural racism should be accelerated.^17^ Bailey, et al, proposed that structural racism may exert its effects through three main routes: residential segregation and its downstream effects on poverty; incarceration; and access to healthcare.^18^ We examined associations in two of these three domains. Our findings support these calls by scientific leaders to focus on structural factors as a part of a national effort to eliminate health inequities. Indeed, past efforts to lessen health disparities in CHD have focused largely on individuals’ physiologic risk factors such as blood pressure and cholesterol and specific health behaviors such as smoking and physical activity. While crucial, structural-level efforts (e.g., expanding Medicaid) may have longer-lasting impact on health inequities with a concomitant focus on more upstream root causes created by our social policies.

To our knowledge, this is the first report of an association between variables reflecting state-level structural racism and the incidence of CHD in a longitudinal cohort. While other studies have reported associations between residential segregation and health outcomes such as cardiovascular health^19^ or incident kidney failure^20^ no prior reports have examined associations between multiple domains of structural racism and clinically adjudicated CHD events longitudinally. A prior study led by Lukachko used 2001-2002 data from the National Epidemiologic Survey on Alcohol and Related Conditions and examined cross-sectional associations between self-report of a physician-confirmed heart attack in the past year and variables representing structural racism in political participation, employment, education, and judicial treatment.^12^ Similar to our study, Lukachko et al showed that Black but not White individuals living in states with high structural racism were more likely to report having a heart attack in the prior year compared to residents of states with low structural racism. A key difference in the approach taken by Lukachko and our study was our examination of health insurance as a domain of structural racism, since social structures result in Black people being limited to low paying jobs without insurance benefits. We found that Black people living in states with higher proportions of Black residents lacking health insurance relative to White residents were at higher risk of incident CHD. Access to health insurance is directly traceable to state-level policies such as the expansion of Medicaid benefits under the Affordable Care Act.^21^ Unlike Lukachko et al, we studied a direct measure of residential segregation by using the DI, which we found to be associated with incident CHD for Black but not White participants. Stricter enforcement of policies and laws intended to expand access to homeownership for Black people could lessen the effects of residential segregation. Results suggest that all three domains of structural racism studied here may act through individual income and education, demonstrating the potential impact of societal structures on socioeconomic resources, which have long been demonstrated to affect health.^22^

Another new finding in our study was the strong association between incident fatal CHD events and structural racism in Black individuals, with no association observed for nonfatal CHD. The strongest association was for health insurance, reflecting the importance of accessing preventive services that can reduce the incidence of fatal CHD. Indeed, our past work has shown that the largest disparities in CHD are for incident fatal CHD.^16^ Most of these events are sudden deaths; risks for sudden death include diabetes, obesity, and cigarette smoking,^23^ all conditions that benefit from healthcare services and all conditions that are more prevalent among Black vs. White individuals.^22^ This study’s numerous strengths include the national cohort, the large sample of Black participants, and the wealth of rigorously assessed characteristics of participants that were available for this analysis. The availability of expert adjudicated incident events is another important strength that adds to the growing evidence that structural racism does have health impact. Limitations include a relatively limited set of structural racism variables and the self-reported nature of some variables that may be prone to recall bias. The observational nature of the study also limits the ability to draw causal inferences.

In conclusion, we report higher risk of incident CHD for Black residents of states with high structural racism on three important domains of social structures: living in poverty, access to health insurance, and residential segregation. These relationships were not evident among White residents of those states. The associations we observed were particularly strong for incident fatal CHD and health insurance. Our findings support the proposition that structural racism contributes to health inequities in incident CHD in the US. State-level policy changes to improve equitable access to income and health insurance and reduce residential segregation should be carefully considered as potential strategies to improve CHD for Black Americans.

## Data Availability

Because of the sensitive nature of the data collected for this study, requests to access REGARDS study data from qualified researchers trained in human subject confidentiality protocols may be sent to the REGARDS study at. The data use agreement with the Centers for Medicare and Medicaid Services does not allow the authors to share participant-level data on Medicare claims. Data management and statistical code is available upon request to the corresponding author.

## Acknowledgements

We thank investigators, staff, and participants of the Reasons for Geographic and Racial Differences in Stroke (REGARDS) study for their valuable contributions. A full list of investigators and institutions can be found at http://www.regardsstudy.org.

## Supplemental Materials

Tables 1-2

Data Sharing Statement

